# Cost-effectiveness of dementia insurance for cognitively-unimpaired *APOE-ε4* homozygotes – a simulation study

**DOI:** 10.1101/2024.11.12.24317164

**Authors:** Kenichiro Sato, Saki Nakashima, Yoshiki Niimi, Takeshi Iwatsubo

## Abstract

Dementia insurance, a private insurance product covering the first diagnosis of dementia of the insured, may be economical for asymptomatic individuals who are aware of their own high genetic risk of developing Alzheimer’s disease (AD) in advance. This is a retrospective study conducted based on National Alzheimer’s Coordinating Center (NACC) data including cognitively unimpaired individuals, aiming to simulate income and expenses of dementia insurance for the insured perspectives. Loss ratio (= total benefits gained / total premium paid) was calculated by *APOE*-ε4 subgroup as a measure of cost-effectiveness, applying the premium rates of actual dementia insurance products being sold in Japan. As a result, for up to 18 years of longitudinal follow-up, the estimated cost-effectiveness improved over the longer observation periods. In individuals in their 60s or older at baseline, the cost-effectiveness was best in the *APOE*-ε4 homozygotes, followed by heterozygotes, and ε4-negative individuals. The dementia insurance for ε4-homozygotes for observation periods ≥ 10 years in this age group was approximately 3 to 4 times more economical than for ε4-negative individuals. Although actively pursuing *APOE* testing for asymptomatic individuals may not be currently recommended due to the concern of adverse selection in the insurance and the absence of available disease-modifying therapy approved for the preclinical stage of AD, our study may provide an important basis for further investigating the advantages and limitations of dementia insurance for asymptomatic individuals with pathogenic or high-risk genes.

## 1 Introduction

Along with the growing population of elderly individuals, there is an increasing awareness among people of the need to prepare for life after retirement, including expenses for long-term care services and medical care services [Yamazaki 2020]. In line with this trend, a private insurance product called “dementia insurance” has been introduced over the past 10-20 years in some countries such as South Korea [Park 2017; RGA HP] and Japan [Šestáková 2019; Goncharova 2020; Eisai HP]. The dementia insurance product is typically designed as diagnostic coverage for middle-aged or elderly non-demented individuals, providing a predetermined insurance benefit if the insured individuals are diagnosed with dementia at certain stages for the first time during the policy period [Eisai HP]. Additional benefits are sometimes provided, such as coverage for dementia prevention, dementia treatment, long-term care services, and medical care services [Park 2017; RGA HP].

In Japan, a super-aging society [Muramatsu 2011] where elderly population comprised 29% in 2021 [eStat], the sale of dementia insurance is reportedly not very popular (e.g., 6.6% of households from a 2021 survey of 4,000 in Japan had purchased it [Japanese Institute of Life Insurance]). This may be due, in part, to the good availability of national health and long-term care (LTC) insurance [Tamiya 2011]. A typical dementia insurance product for older individuals (e.g., aged 60 years) requires the insured to pay thousands of JPY as a monthly premium (c.f., USD 1 = JPY 160 in early July 2024) for life, with a benefit amount of JPY 1,000,000 (i.e., USD 6,250 when 1 USD = JPY 160) upon the occurrence of an event (e.g., first diagnosis of dementia and/or certification as requiring nursing care in the long-term care insurance system).

If a person knew that they had an objectively high risk of developing dementia in the future, they might consider it beneficial for them to purchase a dementia insurance policy while still cognitively unimpaired. However, such objective information is usually not available. For this reason, dementia insurance might be viewed as a viable insurance product for private insurance companies that need to make a profit.

Meanwhile, in some rare cases, people have been found to have an objectively high risk of developing AD, which is the most common cause of dementia [Jack 2013]. These include those with familial AD mutations or two *APOE*-ε4 alleles [Liu 2013; Nicolas 2016; Husain 2021; Fortea 2023]. *APOE*-ε4 homozygotes are at very high risk of developing AD (e.g., odds ratio (OR) 5.9 in ε4/ε3 and OR 33.1 in ε4/ε4 compared to ε4-negative in Japanese population [Farrer 1997]), and they are more prevalent than those with familial AD mutations in the general population [Nicolas 2016]. While *APOE* genotype testing has not been covered by health insurance [Arias 2021], it may become more common testing with the recent approval of anti-amyloid therapies (such as lecanemab [van Dyck 2023] and donanemab [Sims 2023]) and the subsequent need to stratify the risk of developing a specific adverse effect called Amyloid-Related Imaging Abnormalities (ARIA) [Sperling 2011]. This, in turn, may lead to an increase in the number of people unexpectedly found to be ε4 homozygous at their asymptomatic stage, when there is no treatment available, apart from the requirement for testing in the context of anti-amyloid treatment.

Currently, private insurance companies in some regions and countries – mainly Western countries – are not allowed to use genetic information in their underwriting process to prevent genetic discrimination [Muto 2023]. For example, in the United States, it is legally prohibited for medical insurance under federal law, and for other kinds of insurance under state law in some states. In Japan, it is self-regulated by insurance industry association based on a voluntary guideline issued in May 2022 [The Life Insurance Association of Japan 2022]. Current underwriting and payment practices for dementia insurance in Japan do not exclude individuals with a genetically high risk of developing AD, regardless of whether they know their objective risk in advance, as long as they are asymptomatic at the time of insurance purchase.

Private insurance products targeting the general population and operated by for-profit companies are, like lotteries, basically loss-making in terms of expected income versus expenses from the viewpoint of the insured. The same can be said for dementia insurance, where it is estimated that more than half of the policyholders would be relatively low-risk, *APOE* ε4-negative people. On the other hand, ε4-homozygotes are especially susceptible to developing AD, as well as experiencing ARIA during anti-amyloid therapy, and specific opportunities of research and trials for this group are gaining recognition [Fortea 2023]. Given the high risk of developing AD in ε4-homozygotes, we hypothesized that there could possibly be a surplus in terms of expected income versus expenses for those who are ε4-homozygotes, or that the dementia insurance may be at least more economical for ε4-homozygotes than for ε4-negative individuals. If so, can dementia insurance possibly be an option for ε4-homozygotes to mitigate their future disease burden?

This study aims to examine the cost-effectiveness of dementia insurance from the perspective of the insured individuals, using long-term, large observational data to emulate the development of AD and dementia within the longitudinal insurance policyholding history. We apply the actual premium rate settings of dementia insurance products sold in Japan to the data, thereby simulating the balance of income and expenses at the genotype group level, and clarifying the degree of cost-effectiveness of dementia insurance for the ε4-homozygotes compared to other genotypes under Japanese circumstances.

## 2 Methods

### 2.1. Study Design

This is a retrospective study using National Alzheimer’s Coordinating Center (NACC) data, a publicly-available database that includes observational data of more than 50,000 unique participants collected from Alzheimer’s Disease Research Centers in the United States [Beekly 2004]. Our study was approved by a local ethics committee [ID:11754-(1)]. Our study requires a sufficient number of participants with two ε4 alleles with a considerable length of observation period, making the NACC data suitable for our simulation despite it being obtained from North American citizens. We applied the actual premium rate settings of dementia insurance products sold in Japan.

### 2.2. Simulation Settings

We regarded the NACC longitudinal observation data of participants, from their baseline to the development of dementia or dropout from the study, as the longitudinal policyholding data of insured individuals. The maximum length of follow-up for NACC participants is approximately 18 years. Let us consider a typical dementia insurance product to simulate: cognitively unimpaired middle-aged or elderly individuals within a certain age range (e.g., 50-75 years old) at enrollment, without significant past medical history that may directly influence cognitive function, are eligible to purchase the insurance. The insured pay a fixed monthly premium until death or policy cancellation, and receive a lump-sum benefit when they are diagnosed with dementia for the first time, regardless of the cause. For simplicity, we set the benefit as JPY 1,000,000 (= USD 6,250 when USD 1 = JPY 160) for this analysis.

We searched the Internet on July 1, 2024, using a Japanese keyword equivalent to “dementia insurance”, and identified nine simplified-issue dementia insurance products with details available online (Table S1). All provided a primary benefit of a one-time diagnostic lump-sum payment, with exclusion period of up to 2 years from the purchase date. The conditions for receiving benefits were similar across products, with many requiring diagnosis of non-treatable dementia by a physician, and some additionally requiring certification of nursing care necessity in the LTC insurance system. Common reasons for denial of insurance coverage included concurrent hospitalization, a history of neuropsychiatric diseases including MCI or dementia, applying for or being certified for public LTC insurance benefits, and recent surgery or hospitalization. The age range for whole-life dementia insurance largely overlapped across products, with a typical minimum age of 40 and maximum of 79.

Among these products, we selected two whole-life dementia insurance products, where the premium rate for diagnostic coverage of dementia development was available by sex and detailed age at enrollment, and where the insurance benefit was lump-sum JPY 1,000,000 without additional plans required to include. The names of these products or companies are not publicized in this study, as reviewing specific products in an identifiable manner is not the aim of this study.

For simulation using NACC data, we included participants whose Clinical Dementia Rating global score (CDR-GS) was 0 at baseline, who had visited study site twice or more, who had no specific medical history (i.e., stroke, Parkinson’s disease, alcohol abuse, schizophrenia, bipolar disorder, depression within two years, traumatic brain injury, and any other neurological diseases) at baseline, who had no dominantly-inherited AD mutations, and whose *APOE* genotype information was available. The event of interest (i.e., first development of dementia) was defined as a CDR-GS of 1 or more for the first time during observation.

### 2.3. Calculation of Income and Expenses

By regarding the NACC longitudinal observation as the insurance policyholding data, we calculated the balance of income and expenses for insured individuals by applying the age-dependent monthly premium data. Figure 1A illustrates several patterns of the period during which monthly premiums were assumed to be paid for up to *T* years (1≤ *r* ≤ 18). Pattern (a) represents those without the occurrence of an event within the period of *T* years, resulting in only premium payments without gaining a benefit. Pattern (b) is for those who developed dementia within the period, and such individuals gain a benefit, having paid premiums until the time of dementia diagnosis. Patterns (c) and (e) resemble the patterns (a) and (b), respectively.

**Figure 1.**
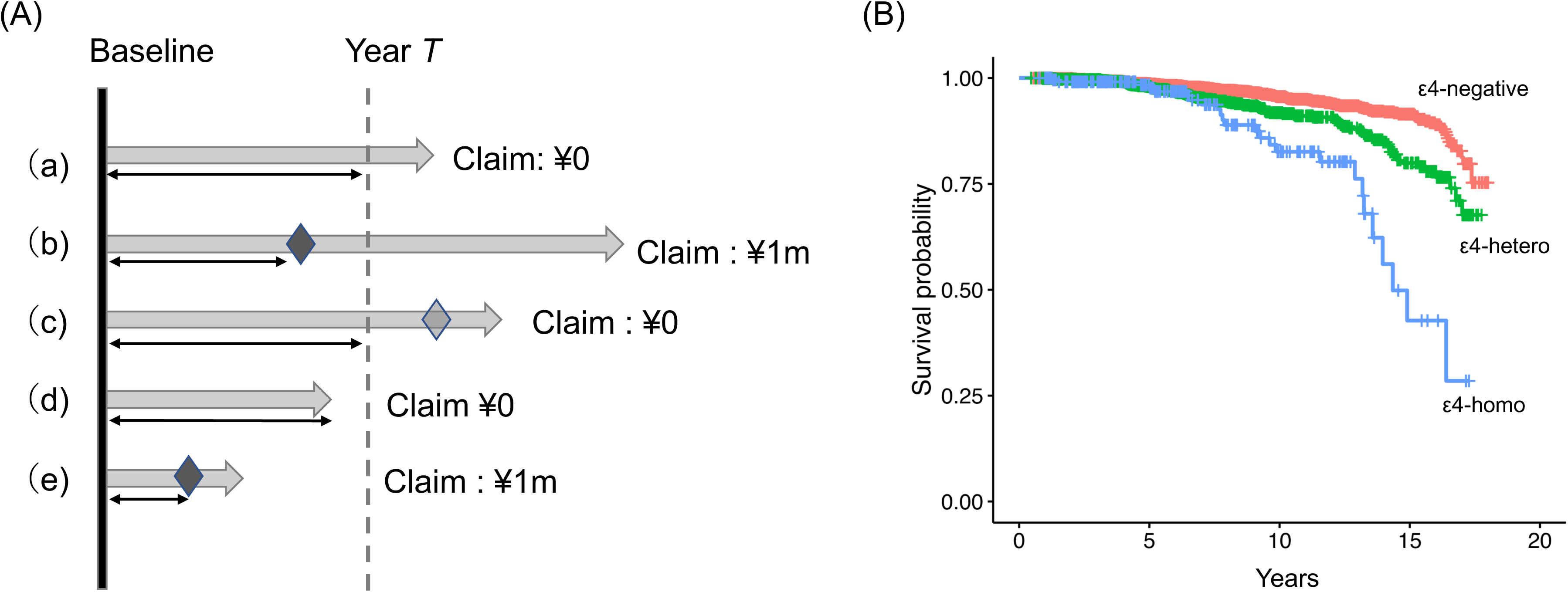
Several patterns in simulating policyholding based on NACC longitudinal observation, and survival curve by *APOE* genotype. In (A), several patterns of the period during which the monthly premium is assumed to be paid for up to *T* years (1≤ *r* ≤ 18) are illustrated. Patterns (a), (c), and (d) represent cases without the occurrence of an event (i.e., development of dementia) within the *T*-year period, while patterns (b) and (e) represent cases where the development of dementia is observed during the period. Diamond shapes in the schema denote the occurrence of the event during the longitudinal observation, and double arrows indicate the period in which the insured are to pay monthly premium. The survival curve of the included participants until their development of dementia (CDR-GS ≥ 1) is shown in (B). Cox-proportional hazards analysis revealed that the hazard ratio (HR) for conversion to CDR-GS ≥ 1 was 1.899 for ε4-heterozygotes and 4.106 for ε4-homozygotes, compared to ε4-negative individuals. Abbreviations: NACC, National Alzheimer’s Coordinating Center; CDR-GS, Clinical Dementia Rating global score.

However, pattern (d) requires attention: individuals in this pattern dropped out of the NACC study without observation of an event during the period, so their income is set to zero. Some of these individuals may have developed dementia after their dropout, meaning their potential income could be underestimated. To address this, we applied two scenarios for balance calculation: scenario (i) assumes pattern (d) individuals canceled their insurance at dropout, while scenario (ii) assumes a fraction of pattern (d) individuals actually developed dementia at dropout. The proportion of those who developed dementia in scenario (ii) is assumed to be the same as those in patterns [b] and [e] combined.

For a given period of *T* years we can obtain the total income from all participants who developed dementia within the period and total payments made by all participants during their policyholding period. This leads to the calculation of loss ratio [Grace 2023], defined as follows:

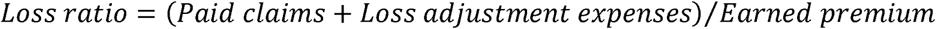

The loss ratio indicates whether an insurance product is financially favorable (< 1) or unfavorable (> 1) for the insurance company. Conversely, a higher loss ratio (> 1) is favorable for the insured, indicating a higher expected value of benefits. The loss adjustment expense (LAE) is the cost incurred by the insurance companies for claim verification. For simplicity, we assumed the LAE to be at most 10% of the total claims. The simplified loss ratio from the insured’s standpoint is defined as:

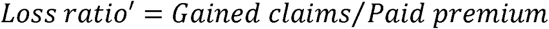

We calculated the loss ratio for each *APOE*-ε4 subgroup (ε4-negative, ε4-heterozygous, and ε4-homozygous). The ratio of the loss ratio between subgroups was further calculated as:

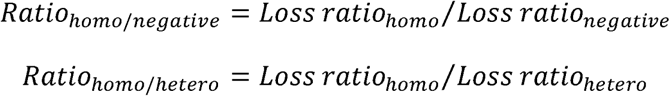

These ratios indicate how much more economical dementia insurance is for the ε4-homozygotes than for the ε4-heterozygotes or ε4-negative individuals. The ratios and their 95% confidence intervals (CIs) are obtained by bootstrapping (B = 1,000). Since younger NACC participants at baseline would be less likely to develop dementia over the 18-year period, we examined the cost-effectiveness metrics for all eligible NACC cases, as well as for the subgroups stratified by the age at baseline (i.e., 50s or younger, 60s, and 70s or older).

## 3 Results

### 3.1. Included Participants

Table 1 shows the baseline characteristics of the examined cognitively unimpaired participants (i.e., CDR-GS of 0) at baseline. The ε4-heterozygotes and homozygotes accounted for approximately 30% and 4%, respectively. An event (i.e., conversion to CDR-GS ≥ 1 for the first time) during the observation period was seen in 3.41% of these participants, with the median occurrence in the 7th year from their observation. The median follow-up period was 4.5 years, with a slight difference in follow-up length by *APOE* genotype (*p = 0.021*, ANOVA test): a Poisson regression model examining the effect of the number of ε4 alleles on follow-up length revealed that ε4-homozygotes and ε4-heterozygotes had 0.900 and 0.968 times shorter maximum follow-up periods compared to ε4-negative individuals.

**Table 1.** Basic characteristics of NACC participants whose age at baseline is 40-79. Abbreviations: NACC, National Alzheimer’s Coordinating Center; IQR, interquartile range; CDR-GS, Clinical Dementia Rating Global Score; CDR-SOB, Clinical Dementia Rating Sum of Boxes; MMSE, Mini-Mental State Examination; PET, positron emission tomography; CSF, cerebrospinal fluid.

| Total N |  | 9,306 |
| --- | --- | --- |
| Follow-up period (years) |  | Median 4.5 (IQR: 1.7 ~ 8.5) |
| Sex (female) |  | 6,074 (65.3%) |
| Age at baseline |  | Median 69 (IQR: 64 ~ 73) |
| Age | 50s or less | 1,280 (13.7%) |
|  | 60s | 3,760 (40.4%) |
|  | 70s | 4,266 (45.8%) |
| APOE-ε4 | Negative | 6,206 (66.7%) |
|  | Heterozygous | 2,759 (29.6%) |
|  | Homozygous | 341 (3.7%) |
| Race | White (vs others) | 7,116 (76.5%) |
|  | Black (vs others) | 1,676 (18.0%) |
| Education years |  | Median 16 (IQR: 14 ~ 18) |
| CDR-GS at baseline (= 0) |  | 9,306 (100%) |
| CDR-SOB at baseline |  | Median 0 (IQR: 0 ~ 0) |
| MMSE at baseline |  | Median 29 (IQR: 29 ~ 30) |
| Became CDR-GS $\geq 1$ within 18 years | | 391 (4.2%) |
| Timing when CDR-GS became $\geq 1$ (years) | | Median 7.7 (IQR: 5.0 ~ 11.3) |
| Amyloid positive (in PET or CSF)* |  | 89 / 371 (24.0%) |

(\*) Amyloid data has been obtained only in a fraction of participants.

Figure 1B shows the survival curve of participants until their development of dementia (CDR-GS ≥ 1). Cox-proportional hazards analysis, including intercept and *APOE*-ε4 alleles as explanatory variables, revealed a hazard ratio (HR) for conversion to CDR-GS ≥ 1 of 1.899 (95%CI: 1.469 ∼ 2.456) for ε4-heterozygotes and 4.106 (95%CI: 2.674 ∼ 6.305) for ε4-homozygotes, compared to ε4-negative individuals.

### 3.2. Loss Ratio by APOE Genotype

Figure 2A shows the loss ratio in each ε4 subgroup, based on the any baseline age of participants, for different observation periods *T* (1 ≤ T ≤ 18). In either insurance product ([a] or [b]), for observation periods ≥ 10 years, the loss ratio was highest in the *APOE*-ε4 homozygotes, followed by heterozygotes and ε4-negative individuals. The loss ratio for ε4-negative individuals remained consistently low, regardless of the examined range of *T*. As *T* increased, the estimated loss ratio notably rose for ε4-homozygotes, irrespective of the scenario assumed ([i] or [ii]). Meanwhile, the loss ratio did not exceed the economically favorable threshold (loss ratio = 1) even for ε4-homozygotes.

**Figure 2.**
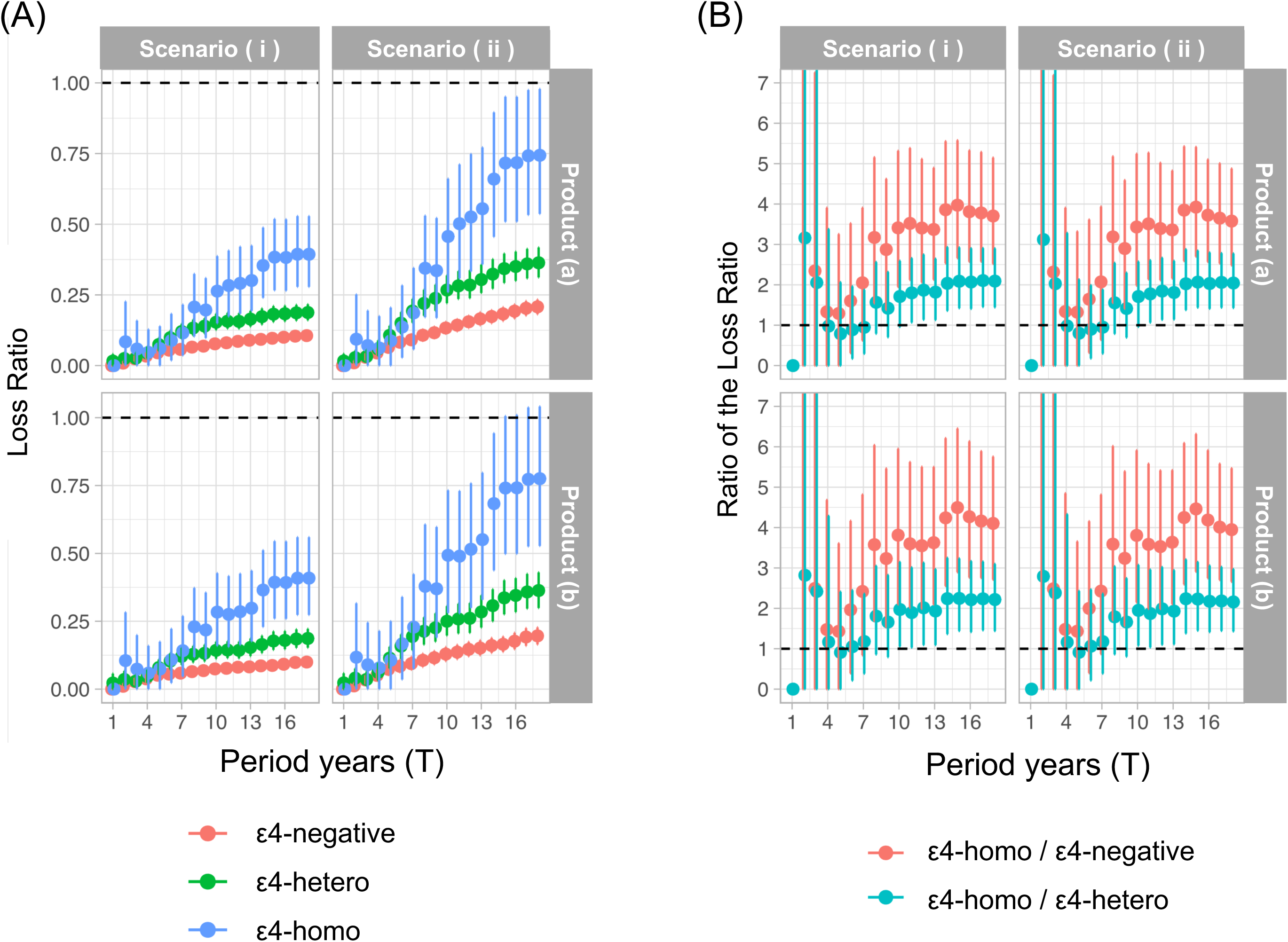
Loss ratio by ε4 subgroup, and the ratio in the loss ratios. In either insurance product ([a] or [b]), for observation periods of ≥ 10 years, the loss ratio (A) was highest in the *APOE*-ε4 homozygotes, followed by heterozygotes, and ε4-negative individuals. The loss ratio for ε4-negative individuals remained low regardless of the examined range of *T*. As *T* increased, the estimated loss ratio rose, particularly for ε4-homozygotes, regardless of whether scenario ([i] or [ii]) was assumed. Meanwhile, the loss ratio did not surpass the threshold of economically favorable level (loss ratio = 1), even for ε4-homozygotes. The ratio of the loss ratios are shown in (B): ratio(homo/hetero) and ratio(homo/negative). Especially broad 95%CIs spanning 1 are observed for T = 2 ∼ 7, and the ratio(homo/negative) reaches as high as 3 ∼ 4, consistently and significantly higher than 1 at T ≥ 8. The ratio(homo/hetero) also displayed a similar pattern, reaching as high as 2, and was consistently and significantly higher than 1 at T ≥ 12. The 95%CIs of the ratio(homo/negative) and the ratio(homo/hetero) show slight overlap with each other.

Figure S1 shows the distribution of loss ratio by baseline age subgroup (50s or younger [Figure S1A], 60s [Figure S1B], and 70s or older [Figure S1C]). Since ε4-homozygotes in their 50s at baseline did not convert to dementia, the loss ratio for individuals in their 50s is zero across all periods (Figure S1A). The loss ratios of individuals in their 60s or older at baseline were similar to the results for all age combined (Figure 2B, C).

### 3.3. Relative Cost-Effectiveness

Figure 2B demonstrates the ratio of the loss ratios: ratio(homo/hetero) and ratio(homo/negative). For T = 1, no meaningful ratios were obtained because no ε4-negative individuals developed dementia within a 1-year period. Broad 95%CIs spanning 1 were observed for T = 2 ∼ 7, but the ratio(homo/negative) consistently and significantly exceeded 1, ranging from 3 to 4 at T ≥ 8. The ratio(homo/hetero) showed a similar distribution, reaching as high as 2 and remaining consistently and significantly above 1 at T ≥ 12. The 95%CIs of the ratio(homo/negative) and the ratio(homo/hetero) showed slight overlap.

Figure S2 presents the ratio of the loss ratios distribution by baseline age subgroup (50s or younger [Figure S2A], 60s [Figure S2B], and 70s or older [Figure S2C]). Since ε4-homozygotes in their 50s at baseline did not reveal conversion to dementia, the ratio for individuals in their 50s at baseline is zero or all periods (Figure S2A). The ratios for individuals in their 70s or older at baseline were similar to the all-age results (Figure S2C), while the ratios for those in their 60s showed limited difference between the ratio(homo/negative) and the ratio(hetero/negative) (Figure S2B).

## 4 Discussion

In this study, we examined the cost-effectiveness of dementia insurance from the perspective of the insured, using longitudinal NACC data to simulate the development of dementia and the subsequent group-level income and expenses. As a result, we found that the estimated loss ratio for ε4-homozygotes increases as the length of the observation period extends, while the loss ratio for ε4-negative individuals remains low regardless of the duration of the period examined (up to 18-year). Furthermore, we identified the loss ratio for ε4-homozygotes is 2 and 3-4 times better than that for e4-heterozygotes and for ε4-negative individuals, respectively, especially among those in their 60s-70s at baseline. These results suggest that non-demented ε4-homozygotes of this age group may find it significantly more economical to purchase dementia insurance than ε4-negative individuals, provided the insurance policy is held for 10 years or longer. Asymptomatic ε4-homozygotes are known as susceptible to developing AD and experiencing ARIA during anti-amyloid therapy, and specific opportunities of research and trials for them are getting recognized [Fortea 2023]. Dementia insurance could potentially serve as a form of support to mitigate their future disease burden. Our results will provide an important basis for further investigating the advantages and shortcomings of dementia insurance for asymptomatic but high-risk people, not limited to *APOE*-ε4 homozygotes.

A major characteristic of this study is applying actual premium rate settings to longitudinal observational data, treating it as the longitudinal insurance history data of the insured. The dementia insurance itself is designed based on the probability of dementia development in the target population; however, there is limited statistical data on the long-term probability of dementia conversion in ε4-homozygotes, as required in our study. This is an advantage of our approach, which overcomes the data shortage.

Meanwhile, simulations using observational data require careful interpretation. NACC participants may be either less or more motivated to continue participating in the study than insured individuals are to maintain an insurance policy. If so, mid-study dropouts from the NACC data may not always be equivalent in frequency to the cancellation of an insurance policy during the period (i.e., scenario [i]). Additionally, the participants’ age significantly influenced the level of cost-effectiveness (Figure S1, S2), due to their difference in the prevalence of dementia development. We were unable to conduct meaningful discussions on the results from participants who were in their 50s or younger (Figure S1A, S2A). Furthermore, although the prevalence of *APOE*-ε4 genotypes in the examined NACC data – *APOE*-ε4-heterozygotes 30% and homozygotes 4% – was not far from real-world estimates, as previously reported [Husain 2021], there is no guarantee that the proportion of ε4-heterozygotes and homozygotes among actual dementia insurance policyholders would be consistent with the prevalence observed in this study. In addition, since the premium rate settings applied were those of dementia insurance sold in Japan, the calculated loss ratio might differ from our results if we had used the premium rate of insurance policies sold in the United States.

For these reasons, the value of the loss ratio itself, as estimated in this study (Figure 2A) for each ε4 genotype, may not be completely reliable. This is also suspect from the aspect of the degree of the loss ratio in ε4-negative individuals (Figure 2A), who should be the majority of the insured: it is lower than 0.25 and appears too favorable for the insurance companies, so the loss ratio estimated in our simulation may actually be underestimated to a considerable degree. This is the major limitation of our study. Although another scenario, assuming that some of the dropout participants had actually developed dementia (i.e., scenario [ii]), attempted to address these shortcomings by demonstrating a larger loss ratio than in the scenario [i], it is necessary to identify the precise method to correct for mid-way dropout cases.

In turn, the relative relationship of the loss ratio between different genotypes, i.e., ratio in the loss ratios (Figure 2B), might be more robust against the shortcomings of directly referring to the loss ratio value. Although this is another measure of cost-effectiveness, it does not directly tell us how economical dementia insurance is, but rather indirectly shows how much more economical dementia insurance is for ε4-homozygotes are than for other ε4 genotypes. This is also easy to understand intuitively, as the ratio in the loss ratios largely corresponds to the risk of developing dementia: the loss ratio for ε4-homozygotes was approximately 3 ∼ 4 and 2 times consistently higher than for ε4-negative and ε4-heterozygous individuals, respectively, and the HR for conversion to CDR-GS ≥ 1 was approximately 2 for ε4-heterozygotes and approximately 4 for ε4-homozygotes. Higher cost-effectiveness here means that ε4-homozygotes will have 3 to 4 times more expected benefits to gain compared to ε4-negative individuals, as well as similar expected benefits with 3 to 4 times lower premium payments than ε4-negative individuals. Based on these evaluations, some ε4-homozygotes in their 60s-70s may find dementia insurance valuable to purchase.

Evaluating dementia insurance as valuable for ε4-homozygotes does not necessarily mean recommending *APOE* testing when considering purchasing dementia insurance, or does it encourage actively pursuing *APOE* testing for asymptomatic individuals as a way to secure insurance with high odds of payoff. Dementia insurance is usually offered only to those without cognitive decline, and *APOE* testing is only considered for individuals who may receive anti-amyloid treatment as part of the risk assessment for developing ARIA [Cummings 2023], with treatment currently limited to MCI or mild AD. This means that people considering purchasing dementia insurance will generally not have access to *APOE* testing in current usual clinical practice. In addition, the prevalence of ε4-homozygote remaining cognitively unimpaired in their 60s-70s is relatively not high: for example, it has been reported that approximately 60% and 40% of cognitively unimpaired ε4-homozygotes in their 60s and 70s, respectively, remain stable over the next 8 years [Bonham 2016]. In these respects, there may be limited situations where our finding will be clearly applicable.

Meanwhile, it is technically possible for ε4-homozygotes to know their ε4 genotype, such as through *APOE* testing services offered in medical practices not covered by health insurance [Sato 2024(a)], via direct-to-consumer services [Pavarini 2021], or by participating in prevention trials or clinical studies related to AD. Indeed, our previous online survey [Sato 2024(b)] revealed that approximately 45% of questionnaire respondents expressed a willingness to undergo *APOE* testing, even without the immediate need for testing prior to anti-amyloid treatment [Sato 2024(c)]. If the potential cost-effectiveness of dementia insurance for ε4-homozygotes becomes widely recognized, it may lead to adverse selection, with ε4-homozygotes being more likely to concentrate in dementia insurance policies far more than the general population, resulting in a significant increase in paid benefits for insurance companies, and eventually necessitating higher premiums or the discontinuation of dementia insurance products in the long term. It may also lead to the misuse of *APOE* testing. Therefore, we do not recommend actively seeking *APOE* testing when considering purchasing dementia insurance. However, it may be unavoidable to consider dementia insurance for exceptional cases, such as those who already know their ε4 status before deciding to purchase insurance. A similarly complex discussion on risk stratification by *APOE* testing prior to purchasing insurance may also apply to blood-based biomarker (BBM) testing results, which now provides highly accurate predictions of pre-symptomatic pathological changes in AD [Niimi 2024], although it is not a genetic test.

In conclusion, our study demonstrated that dementia insurance may be more cost-effective for *APOE*-ε4 homozygotes compared to other genotypes. Although the dementia insurance could serve as a potential supportive option to alleviate the future potential disease burden for some ε4-homozygotes, we would need further investigation on the advantages and limitations of dementia insurance for asymptomatic individuals with pathogenic or high-risk genes, including but not limited to *APOE*-ε4 homozygotes.

## 6 Author Contributions

KS: Conceptualization, Study Design, Data Acquisition, Data Curation, Analysis, Writing draft. SN: Data Acquisition, Review & Editing. YN: Review & Editing. TI: Supervision. All authors read and approved the final manuscript.

## Data Availability

All data produced are available online at National Alzheimer's Coordinating Center (NACC) (https://naccdata.org).

---

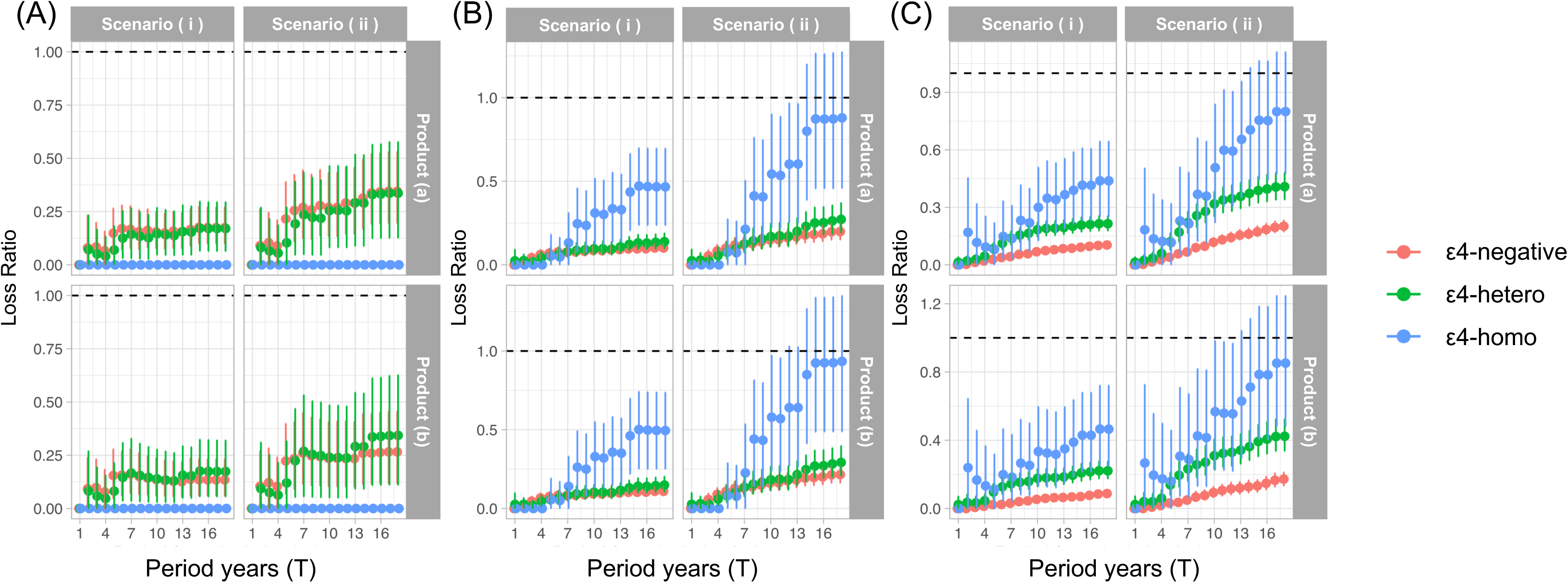

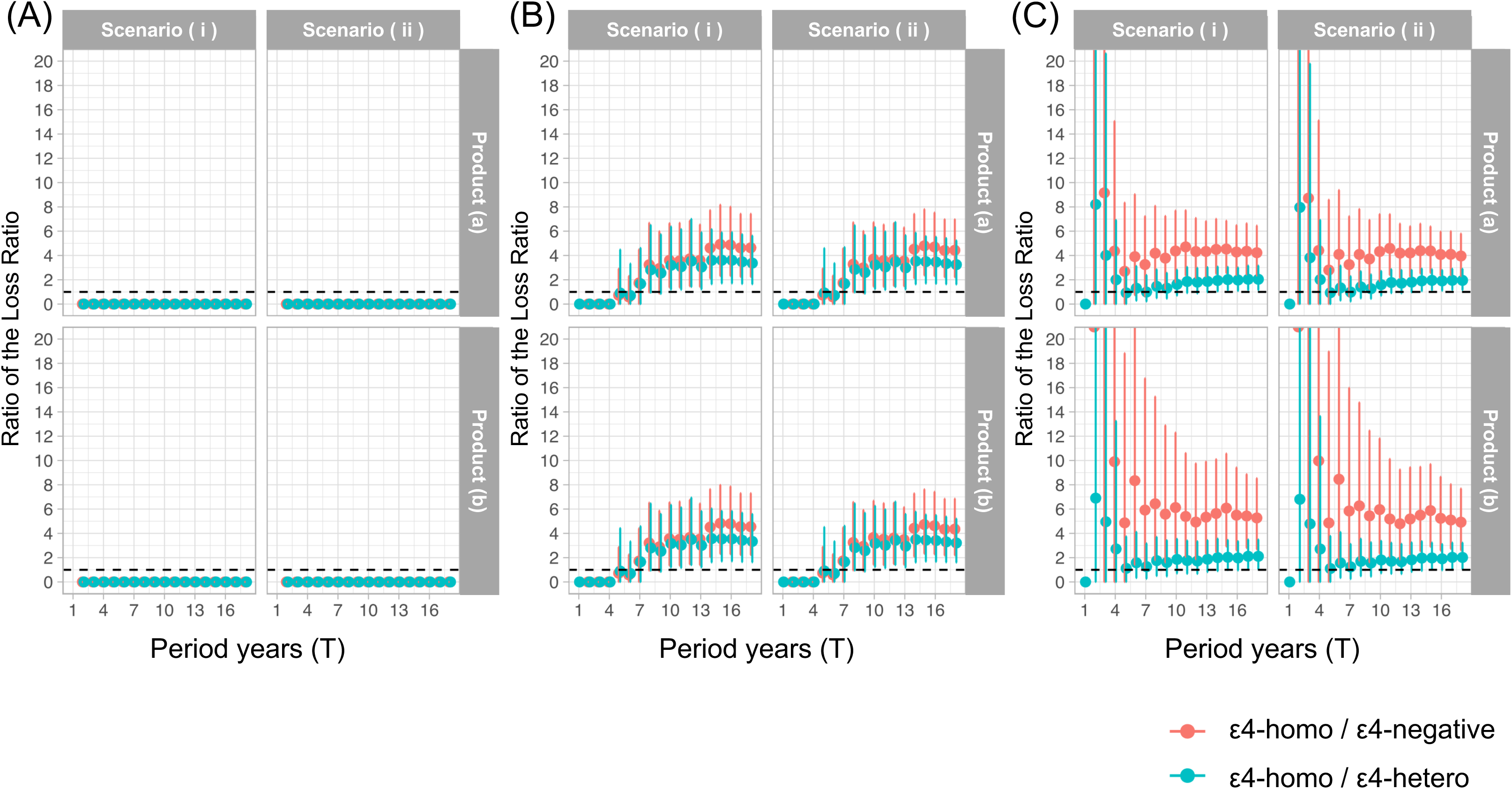

