## Supplementary material for "Cost-effectiveness of dementia insurance for cognitively-unimpaired *APOE-ε4* homozygotes – a simulation study": Table S1

**Supplementary Matrials**

Table S1. List of dementia insurance products sold in Japan we identified by internet search in 2024

| Product # | Conditions of insurance coverage | Exclusion period from the date of purchase of insurance |
| --- | --- | --- |
| #1 | - Diagnosed with prespecified dementia   and   - Certified as LTC nursing care degree 1 | 2 years |
| #2 | - Diagnosed with prespecified dementia | <1 year |
| #3 | - Diagnosed with non-treatable dementia | <1 year |
| #4 | - Diagnosed with non-treatable dementia | 1 year |
| #5 | - Diagnosed with non-treatable dementia | 1 year |
| #6 | - Diagnosed with non-treatable dementia | <1 year |
| #7 | - Diagnosed with prespecified dementia | 1 year |
| #8 | - Diagnosed with non-treatable dementia | 2 years |
| #9 | - Diagnosed with non-treatable dementia   and   - Certified as LTC nursing care degree 1 | 2 years |

Among these insurance products, we selected two to use their premium rate settings (not specified in the above list to avoid identifiability).

Abbreviations: LTC, long-term care.
